# A functional neuroimaging biomarker of mild cognitive impairment using TD-fNIRS

**DOI:** 10.1101/2024.11.06.24316775

**Authors:** Julien Dubois, John Gregory Duffy, Ryan M. Field, Erin M. Koch, Zahra M. Aghajan, Naomi Miller, Katherine L. Perdue, Gregory Sahagian, Moriah Taylor

## Abstract

**INTRODUCTION:** Diagnostic assessments of mild cognitive impairment (MCI) are lengthy and burdensome, highlighting the need for new tools to detect MCI. Time-domain functional near-infrared spectroscopy (TD-fNIRS) can measure brain function in clinical settings and may address this need.

**METHODS:** MCI patients (n=50) and age-matched healthy controls (HC; n=51) underwent TD-fNIRS recordings during cognitive tasks (verbal fluency, N-back). Machine learning models were trained to distinguish MCI from HC using neural activity, cognitive task behavior, and self-reported impairment as input features.

**RESULTS:** Significant group-level differences (MCI vs HC) were demonstrated in self-report, N-back and verbal fluency behavior, and task-related brain activation. Classifier performance was similar when using self-report (AUC=0.76) and self-report plus behavior (AUC=0.79) as input features, but was strongest when neural metrics were included (AUC=0.92).

**DISCUSSION:** This study demonstrates the potential of TD-fNIRS to assess MCI with short brain scans in clinical settings.

## 1. Background

Mild Cognitive Impairment (MCI) is often regarded as the intermediary stage between normal aging and dementia [1]. In fact, 12-18% of older adults (age > 60 years) live with MCI, and of those, 10-15% progress to dementia each year [2]. The emotional and socioeconomic burden of cognitive impairment [3] has been driving the recent advances in developing biomarkers capable of early detection and objective diagnosis of dementia with varying degrees of accuracy[1,4,5]. Early diagnosis could afford patients the opportunity to preserve cognition and halt further progression of the disease. For example, 40% of dementia cases can be avoided with lifestyle adjustments [6]. Furthermore, currently there are disease modifying therapies capable of slowing cognitive decline [5]. Standard assessments are commonly used to screen for potential cognitive decline and refer patients for further testing [7–9]. Diagnostic tools, however, span a range of modalities, including those readily available such as neuropsychological assessments [10] and digital biomarkers [11], as well as those that are still being explored for accessibility and clinical potential, such as neural measurements [12–14].

Many of the commonly used screening tests such as Mini-Mental State Examination (MMSE), and Montreal Cognitive Assessment (MoCA) both fail to identify patients, including those with high cognitive reserve (false negative), but they also incorrectly flag patients with low education and/or high anxiety (false positives) [15]. Flagged patients are further subjected to comprehensive neuropsychiatric assessments which are cumbersome, stressful, and time-consuming for the patient and clinician alike, as they may take up to 3-hours [10]. This burden is exacerbated by the high prevalence of false positives in screening. Further complicating MCI diagnosis is the fact that even in healthy aging, seniors have a high-likelihood of failing at least one memory test, necessitating more than one assessment in that domain [10].

One explanation for these challenges is that neuropsychiatric assessments only measure symptoms of the disease (e.g., cognitive deficits across multiple domains of functioning), while other medical conditions are diagnosed by directly measuring and/or imaging underlying biological substrates of the disease. This explanation suggests that more direct measures of the etiology underlying MCI—specifically, functional brain activity—could offer a more reliable basis for diagnostics. Neural measurements may even predate measurable cognitive deficits and differ when cognitive scores do not. This in turn may negate the need for redundant task administration and excessive probing, significantly reducing the time and burden of the diagnostic protocols currently employed.

Among brain-imaging techniques for MCI diagnosis, functional magnetic resonance imaging (fMRI) has been researched extensively [12,16,17]. Although useful for research purposes, this method has had limited success when it comes to clinical use—in particular in multi-center efforts [16]. Functional near infrared spectroscopy (fNIRS), another brain-imaging tool that uses light to measure the brain and is sensitive to relative oxygenation fluctuations, has emerged as a viable alternative. Dozens of studies over the past 20 years have confirmed that patients diagnosed with MCI have changes in hemodynamic signals detectable by fNIRS measurements (for reviews see [18,19]) As such, multiple studies have explored the potential of fNIRS measurements in MCI diagnosis [20–22]. Time-domain fNIRS (TD-fNIRS) is a more advanced form of fNIRS which has better sensitivity to brain activations [23,24]. Despite these advantages, this technology has not been widely used in clinical trials or clinical care due to the high cost, operational complexity, and limited head coverage of the currently available systems [23].

Here we employ a validated, state-of-the-art TD-fNIRS system with miniaturized components, dense whole head coverage, and the form factor of a helmet [25–27]. The technological advancements of this device greatly improve the portability and ease-of-use, making in-clinic brain imaging accessible for clinical MCI care. In light of this, we assessed the potential of diagnostic brain imaging as a replacement for standard neuropsychiatric evaluations in MCI. Specifically, MCI patients and age-matched healthy-controls (HCs) completed common clinical surveys and short cognitive tasks (< 10 min each) administered on a computer, while their brain data was recorded with the aforementioned TD-fNIRS headset. Survey scores, task performance, and measures of brain activity were extracted, a subset of which were identified as statistically differentiating between the MCI and HC cohorts. These metrics were then used as an input to a series of classification models that quantified the ability of different combinations of each class (e.g. survey, behavioral, and neural metrics) to identify MCI. We show that neural and behavioral metrics offer diagnostic capabilities beyond the current standard of care. Moreover, we present a brain-based biomarker of MCI that is objective, scalable, and fast to administer, highlighting the potential of TD-fNIRS to alleviate the diagnostic and care management burden of this disease on clinicians, patients, and their families.

## 2. Methods

### Participants and screening procedures

Clinically diagnosed MCI patients (n=50) and age-matched healthy controls (HC; n=51) completed this study (NCT05996575). All patients were diagnosed by an expert clinician (neurologist or psychiatrist). Participants gave written informed consent before beginning the study in accordance with the ethical review of the Advarra IRB (#Pro00071712), which approved this study, and the Declaration of Helsinki. Patient data was collected at two clinical locations, and healthy control data was collected at both clinical locations and Kernel.

All participants were required to meet the following inclusion criteria: (1) 55–85 years of age, inclusive at time of enrollment, (2) the ability to perform informed consent on their own, (3) fluent in English (speaking and reading), and none of the following exclusion criteria: (1) current substance or alcohol dependence, and/or alcohol or substance abuse as determined by CAGE-AID assessment for drug and alcohol abuse, with a score of 2 or higher resulting in exclusion, (2) uncorrected major visual or auditory deficits that would prevent them from completing a study task, (3) current or recent (in the past 6 months) chemotherapy and/or radiation for any cancer, and (4) major medical illnesses and psychiatric conditions (other than MCI). Additionally, MCI patients were eligible to participate in the study if they had a diagnosis of MCI (amnesiac or non-amnesiac) as determined by clinician, and did not have an Alzheimer’s or dementia diagnosis. Moreover, healthy controls were eligible to participate in the study if they fulfilled none of the following additional exclusion criteria: (1) prior MCI or memory impairment diagnosis, (2) first-degree relative with dementia or clinically relevant memory problems, and (3) Alzheimer’s or dementia diagnosis.

### Study design

#### General description

All three tasks were designed and presented using the Unity game engine (Figure 1).

**Figure 1.**
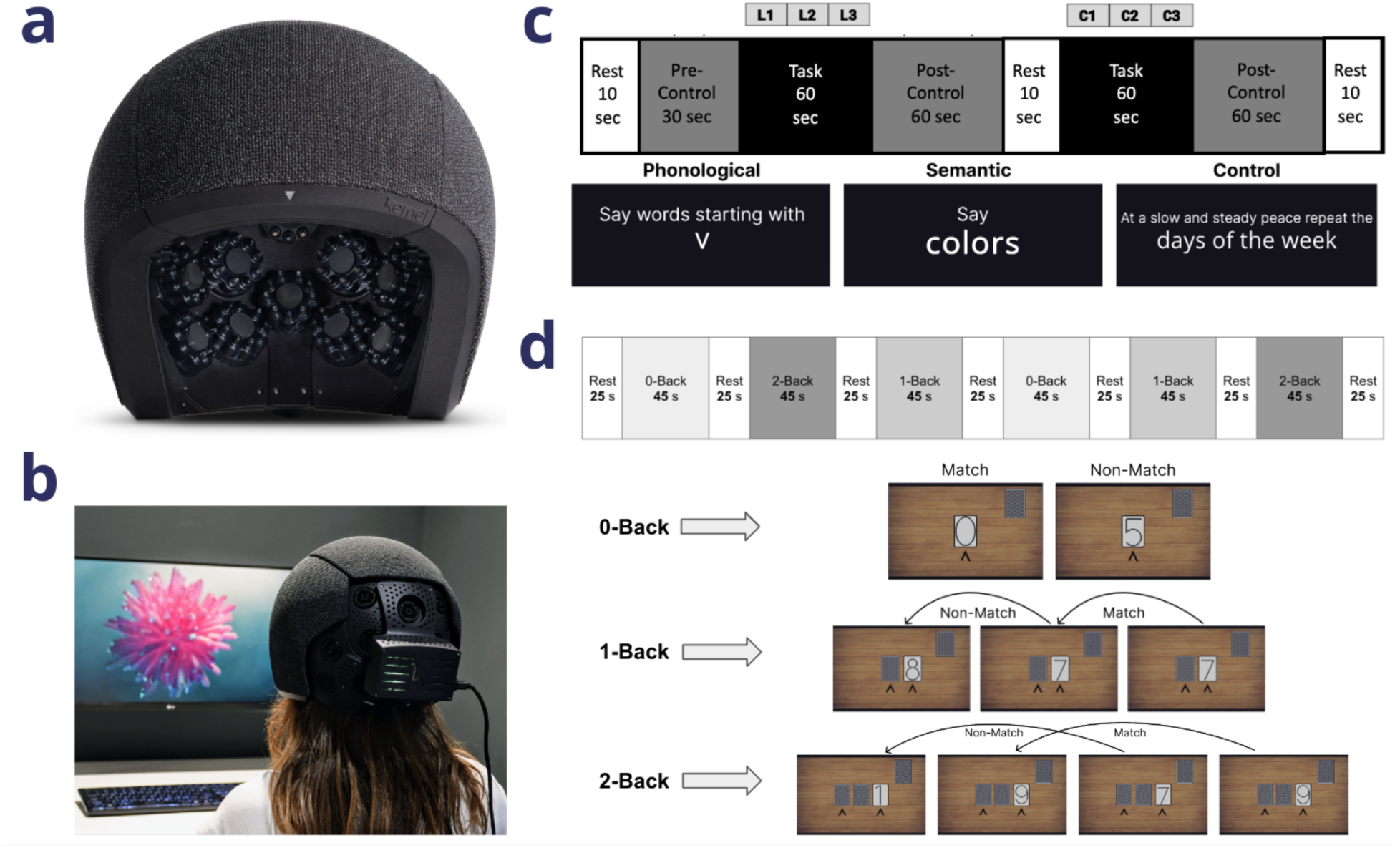
Overview of the experimental setup. **a.** The Kernel Flow2 TD-fNIRS headset, **b.** Model demonstrating wearing the headset while brain data is recorded during a resting state session, **c.** Verbal Fluency Task design, **d.** N-Back task design.

#### Resting State

During this task, participants watched a 7-minute audiovisual clip without any other particular task demands[28] (Figure 1b). Data from this task was not used for analyses in this manuscript as here we focused on cognitive tasks that probe domains currently included in clinical assessments.

#### VFT task paradigm

In the Verbal Fluency task (Figure 1c), participants orally produced words in response to three types of prompts: phonological (words beginning with a specified letter), semantic (words from a given category), and control (reciting the days of the week repeatedly). Prompts appeared in white text on a black screen. Control instructions read, “At a slow and steady pace, repeat the days of the week.” Phonological prompts read, “Say words starting with {insert letter},” and semantic prompts read, “Say {insert category}.” Participants were instructed to generate as many relevant words as possible for as long as each prompt was on the screen. They were instructed to avoid repeat words and proper nouns. They were also required to press the spacebar each time they produced a word. They were told that they could whisper or speak the words aloud.

The task began with a 10-second rest period (reminding participants to press the spacebar) followed by a 20-second control block (“precontrol“; not utilized in behavioral or neural analysis) to get the participant in the rhythm of the task. It then progressed through a 60-second phonological block, in which 3 different letters were presented in succession for 20 seconds each. A 60-second control block followed. After a 10-second rest, the semantic block began, with three categories presented for 20 seconds each. The final block was a 60-second control block, and the task concluded with a 10 second rest period.

All participants received identical prompts in the same order. The phonological prompts were F/A/S, and the semantic prompts were animals/fruits/sports. Before data collection, participants performed a practice session during which a semantic prompt of “colors” and phonological prompt of letter “V” was used.

#### N-back task paradigm

The N-Back task is a working memory task where participants responded when the presented stimulus matched one presented N steps earlier (Figure 1d). The difficulty was adjusted by modifying the load factor “N” (Figure 1b). In this implementation, the task was gamified to resemble a card game for increased engagement. It included three block types: 0-back, 1-back, and 2-back, using numbers displayed on virtual playing cards. All blocks of the task began with a stack of face-down cards in the upper right corner of the screen.

In 0-back blocks, one card at a time slid down from the stack to the center of the screen, flipped to reveal its number, remained face-up for 0.75 seconds, and then flipped face-down as it slid left and exited the screen. Participants were asked to press the spacebar as quickly as possible when a card with the number “0” appeared face-up. This repeated for 28 trials.

In 1-back blocks, participants were asked to press the spacebar if the number on the current card matched the one flipped immediately before it. The card movement was similar to a 0-back block, but as each card flipped face-down it slid to the left and remained face-down on the screen as the next card appeared and flipped face-up. If the numbers on two consecutive cards (the face-up and face-down cards) matched, the participant was instructed to press the spacebar. This process also repeated for 28 trials.

Finally, in 2-back blocks, participants were asked to press the spacebar if the number on the current card matched the number presented two cards prior. The motion was similar to a 1-back block, but three consecutive cards remained on the screen at once—two facedown on the left and the current card face-up on the right. If the leftmost card matched the face-up card, the participant was to press the spacebar. The leftmost card slid off the screen when a new card appeared. This repeated for 28 trials.

Each task run generated unique stimuli with a 25% match rate. The block order was always 0-back, 2-back, 1-back, 0-back, 1-back, 2-back. A 25-second rest period was included between blocks and at the beginning and end of the task. Prior to data collection, each participant completed a guided tutorial combining instructions and practice.

### Cognitive screening and survey data collection

Multiple surveys were administered to the participants during the study. For the HC population, the Mini-Cog [29] was employed as a quick assessment tool for dementia screening. With the scores varying between 0 and 5, a cutoff of >=3 was used to verify healthy functioning, as is customary. For the MCI population, the Mini-Mental State Examination (MMSE) [8,30] scores were collected from the participating clinics. Here, the range of the scores is 0-30, with lower scores indicating lower cognitive functioning. There were additional surveys that were administered to both HC and MCI cohorts:

1. Self-reported Alzheimer’s Disease Cooperative Study Activities of Daily Living (ADL) for Mild Cognitive Impairment (ADCS-ADL-MCI) survey [31]: This survey, which aims at assessing the degree of functional impairment as a result of cognitive impairment, consists of 20 questions and results in a score between 0-49, with lower scores suggesting more functional/cognitive impairment.
2. General Anxiety Disorder (GAD)-7 [32]: Commonly used as an initial screening tool for screening anxiety disorders, this questionnaire consists of 7 questions, resulting in a score between 0-21, where higher scores suggest more severe symptoms.
3. Geriatric Depression Scale (GDS) [33]: In older adults, this 15-item self-report questionnaire is an initial screening tool for depression. Higher scores indicate higher symptom severity.

### fNIRS data collection and feature extraction

#### Data acquisition

Neurophysiological data were collected using the Kernel Flow2 TD-fNIRS [26,27] as participants wore the headset during cognitive tasks and resting state sessions. Systems specifications were described in detail previously [25]. Briefly, distributions of the times of flight of photons (DTOFs) of over 3000 source-detector pairs (channels) from 40 modules arranged over the prefrontal, parietal, temporal, and occipital areas were recorded (effective sampling rate 3.76Hz) (Figure 1a). Of the original configuration of modules, 35 were deemed to be most relevant to our tasks (the most posterior modules over the inferior occipital regions were removed)(Supplement S1). Note that the L/R parietal regions spans mid to fronto-parietal cortex, while the occipital region spans occipital to posterior parietal cortex.

#### Data preprocessing and relative hemoglobin concentrations

Data preprocessing was done similar to those in prior publications [25,34,35]. A channel selection procedure (using histogram shape) [27] was followed by computing DTOFs’ moments (i.e., sum, mean, and variance moments) [36,37]. We then used a sensitivity method [24], deriving sensitivities from a finite element modeling forward model from NIRFAST [38,39], to obtain the relative changes in absorption coefficients for each wavelength, which were subsequently converted to changes in oxyhemoglobin and deoxyhemoglobin concentrations (HbO and HbR, respectively) using the the modified Beer–Lambert law (mBLL)[40]. These signals were further processed using motion correction (Temporal Derivative Distribution Repair[41]), cubic spline interpolation [42], detrending using a moving average algorithm (100-second kernel), and finally short channel regression[43] (SDS=8mm).

#### Participant Exclusion

Data was initially collected from 130 participants, 65 HCs and 65 MCI participants. One participant was withdrawn from the HC group after data collection due to being below the age inclusion limit of 55 (screen fail). Three participants were withdrawn from the MCI group: 1 due to being above the age inclusion limit of 85 (screen fail) and 2 due to technical difficulties preventing data collection. This left 64 participants in the HC group and 62 in the MCI group. Of those in the HC group, 8 were excluded from the data analysis who were between the ages of 55 and 59. This decision was made to age-match to the MCI group. Further, only participants with complete datasets, i.e., participants who finished the entire protocol were considered for further analysis. Of the 56 remaining participants in the HC group, 2 healthy controls completed an outdated version of the N-back task which had mistakenly been used, 1 encountered hardware issues, 1 failed to complete resting state due to head discomfort, and 1 had abnormally low signal due to operator’s failure to follow proper headset set-up procedures. Therefore, 5 participants had only partial datasets (missing 1 or more of the 3 tasks), and 51 HC’s were included in the analyses presented here. Of the 62 participants in the MCI group, 12 had only partial datasets. Reasons for the 12 incomplete datasets are as follows: 1 completed an outdated version of N-back, 1 stopped N-back early due to discomfort, 2 faced technical issues loading the resting state video, 2 encountered wifi issues that prevented data collection, 1 participant fell asleep, 3 had unexplained cropped session recordings, and 2 encountered hardware issues. This left 50 MCI participants with all three task sessions, which were included in analyses.

#### General linear model (GLM) approach

As described previously in detail [35], a design matrix (X: a set of task specific regressors), and residuals (*ε*) were used to model the time course of each channel (y: relative HbO and separately relative HbR) using the following equation: y = X*β* + *ε*. Here, the *β* coefficients represent the contribution of each regressor. A least-squares solution combined with prewhitening of the time courses (using an autoregressive model of order 15, i.e., 4 × the sampling rate of our system) was used to solve this multiple linear regression problem. To construct the design matrix, X, we used the following regressors: (1) time course of the task blocks as square waves that were then convolved with a canonical hemodynamic response function for each block type); (2) drift regressors; and (3) a standard set of Discrete Cosine Transform regressors to remove trends in the data (with a period of 100s and more) as implemented in nilearn Python package[44]. Test statistics, commonly referred to as the GLM Contrasts, and their p-values were computed between conditions of interest (using t-tests) on the *β* coefficients. This approach was used for both the Verbal fluency and N-Back tasks.

1. *<u>Verbal Fluency task</u>*: The three block types in the experimental design, i.e., control, semantic, and phonological were used as GLM contrasts..
2. *<u>N-Back task</u>*: Similar to the verbal fluency task, the three block types in the experimental design, i.e., 0-back, 1-back, and 2-back were used as GLM contrasts.

### Machine learning (ML) for classification

We used a logistic regression model to predict whether a participant belonged to the HC or MCI group (Figure 2; Supplement S2). Expert clinician diagnosis was used to label MCI rather than the score on any particular survey, cognitive test, or questionnaire. We trained and tested different models depending on the inputs (described below). Stratified nested K-fold cross validation was used (N_inner_ = 10, N_outer_=10) where the inner loop was used for regressing out confounds, feature selection, and hyperparameter optimization, while the outer loop was used to obtain the model performance on the independent test sets. An elastic net regularization with L1-ratio=0.5 was used to push feature weights towards zero and avoid overfitting with a large number of input features.

**Figure 2.**
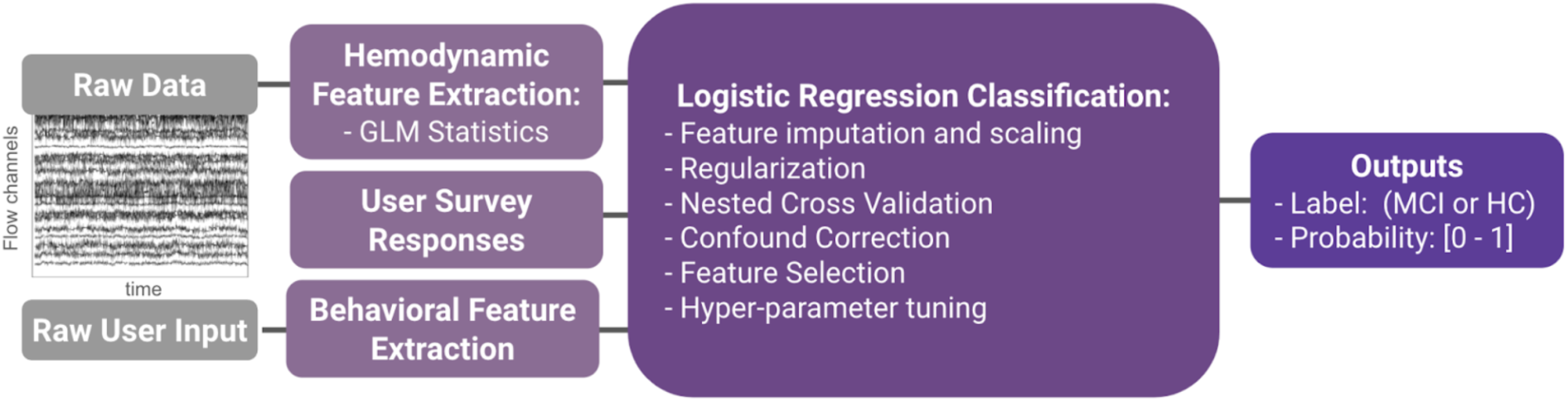
Overall structure of the machine learning model for MCI classification.

In order to prevent data leakage, the following steps were fit on the training set of the inner cross-validation loop and applied to the test set:

1. *Imputation of input data*: missing input data, which could result from poor headset coupling in a region, was filled in as the mean value from the training set population.
2. *Standardization of the input data*: Each input feature was z-scored with respect to the training set population.
3. *Residualization of confound variables*: Age and gender both exhibited relationships with input features and thus were regressed out from the input features. Specifically, a linear model was fit between the confound variables and each input feature. Then the residuals were computed and replaced the original input features, as they represent the variability in each metric that could not be explained by the confound variables.
4. *Feature selection*: Feature selection was implemented using statistical tests, namely by applying independent t-tests on a given feature between the two HC and MCI cohorts. The p-value from this test was later used in step (5).
5. *Hyperparameter Tuning*: The only hyperparameter that was tuned for this model was the alpha threshold, which was allowed to vary between 0.05-0.30 in steps of 0.05.

On each outer fold, the best hyperparameter from the inner loop cross validation was used. The model yielded both a predicted label (binary output) and a raw score (probability between 0 and 1). Predictions and scores were concatenated across all outer loops, resulting in one prediction for each participant.

To benchmark the performance of different types of input features we fit 3 different models: one using only survey response (score from ADCS-ADL-MCI) as input, one using survey responses and behavioral metrics from the two cognitive tasks as input, and one adding neural data thus yielding survey, behavioral, and brain metrics as input. Importantly, the folds were fixed such that they were the same when training all three models, so results could be directly compared. We also trained a model limiting the inputs to those available during the cognitive tasks, i.e., brain and behavioral metrics to investigate the necessity of the survey data to model performance.

To determine which features were contributing the most to the model, we ranked features by the absolute value of the average feature weights over the outer folds of the model and visualized the top 50 input features.

We computed the following metrics to capture the performance of our models in detecting MCI:

1. *Confusion matrix*: computing the number of true positives (TP), true negatives (TN), false positives (FP) and false negatives (FN)
2. *Accuracy*: (TN + TP)/N_total_
3. *Recall (Sensitivity)*: TP/(TP + FN)
4. *Precision*: TP/(TP + FP)
5. *Specificity*: TN/(TN + FP)
6. *Negative Predictive Value (NPV)*: TN/(TN + FN)
7. *AUC*: Area under the receiver operating characteristic (ROC) curve.
8. *Adaptive thresholding of the model:* Here we followed techniques demonstrated in the literature [45] to split the model’s decision threshold into 2, creating a new label where scores between these thresholds are deemed “Inconclusive” or “Flagged for a “Follow-up”. Specifically, the upper threshold is adjusted such that it is the lowest value satisfying a particular specificity (here 0.90) and then the bottom threshold is similarly adjusted to be the highest value satisfying a particular sensitivity (here 0.90).

## 3. Results

### General description

The two participant groups, MCI and age-matched HC (Table 1) wore the Kernel Flow2 TD-fNIRS headset (Figure 1a) and their neurophysiological data was measured while they performed three tasks: resting state (Figure 1b), verbal fluency (Figure 1c) and N-back working memory (Figure 1d). We verified that the two populations were truly age-matched by performing an independent t-test (p>0.1). The groups were similar in terms of the number of participants with higher education as well (Table 1).

**Table 1.** Participant demographics and characteristics. For the survey scores, shown are the mean±standard deviation.

|  | HC | MCI |
| --- | --- | --- |
| **Number of participants** | 51 | 50 |
| **Age** | 71.39 ± 6.87 | 73.68 ± 6.49 |
| **Gender**<br>*Female*<br>*Male* | 35<br>16 | 30<br>20 |
| **Higher Education**<br>*No degree*<br>*Associate's degree*<br>*Bachelor's degree*<br>*Postgraduate/Professional degree* | 13<br>10<br>16<br>12 | 19<br>8<br>12<br>11 |
| **Race**<br>*American Indian/Alaska Native*<br>*Asian*<br>*Black or African American*<br>*Native Hawaiian/Pacific Islander*<br>*White/Caucasian*<br>*Unreported* | 1<br>2<br>6<br>0<br>41<br>1 | 1<br>3<br>2<br>1<br>43<br>0 |
| **Ethnicity**<br>*Hispanic, Latino/a, and/or Spanish origin*<br>*Unreported* | 5<br>1 | 5<br>1 |
| **Tobacco Use**<br>*Current or former smoker* | 19 | 26 |
| **MMSE (range: 0–30)** | NA | 26.30 ± 2.43 |
| **Mini-Cog (range: 0–5)** | 4.59 ± 0.63 | NA |
| **ADCS-ADL-MCI (range: 0–49)** | 42.90 ± 2.90 | 37.71 ± 7.13 |
| **GDS (range: 0–15)** | 1.31 ± 1.97 | 2.46 ± 2.23 |
| **GAD-7 (range: 0–21)** | 1.76 ± 2.50 | 2.90 ± 3.44 |

Our analyses, discussed below, focused first on characterizing the populations in terms of their cognitive assessment and survey scores. Next, we sought to find cognitive task-based features, both in terms of behavioral and neural measures that significantly differentiated between the two study cohorts. Finally, we aimed at building an ML model capable of classifying MCI with high performance.

### Participant cognitive assessment and survey results

Cognitive assessments were performed in both the HC and MCI groups (Methods). In the HC population, this included the Mini-Cog, for which the majority of the participants had a score of 5 (Figure 3a). In the MCI population, we examined the MMSE scores. Although many of the participants had a score above 25 (Figure 3b), a cutoff often considered as normal cognitive function, the true clinical diagnosis was determined by expert clinicians (using a variety of tools).. In fact, it has been shown that with a cutoff score of 25 (inclusive), MMSE sensitivity for MCI detection is only 18% [7,46]. The only cognitive assessment done with both populations was the ADCS-ADL-MCI survey (Methods). As expected, survey scores were significantly different between MCI and HC (t-test p=5.43×10^-6^) with the MCI group reporting more functional impairment (lower scores; Figure 3c).

**Figure 3.**
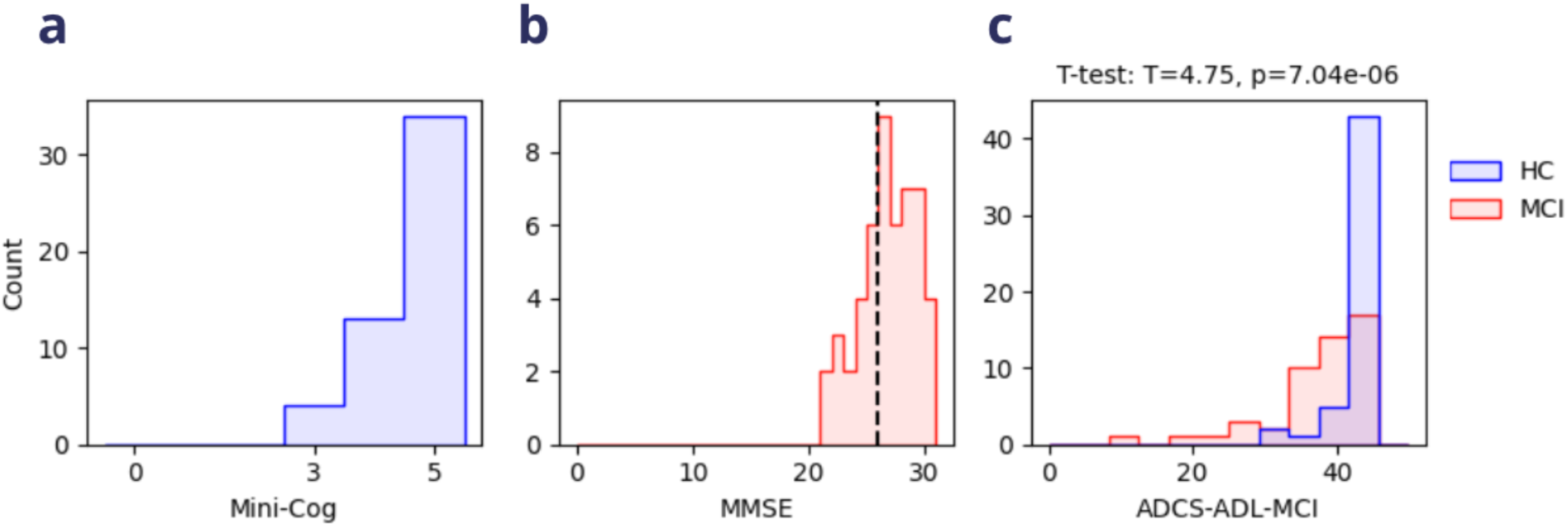
Participants’ survey scores in the HC and MCI groups. **a.** The distribution of the Mini-cog test scores, which was administered only to the HC population. The range of the scores is 0-5, with 5 indicating no cognitive decline. HC inclusion required a score>=3. b. The distribution of the MMSE scores in the MCI population. The range of the scores is 0-30, with scores<=25 suggesting cognitive impairment (dashed line). There was no threshold for inclusion, c. The ADCS-ADL-MCI survey scores were significantly different between the HC (blue) and MCI (red) populations with higher scores indicating better daily functioning.

We examined the other two non-cognitive surveys, i.e., GAD-7 and GDS (Methods). GAD-7 showed a trend towards higher anxiety scores in the MCI population (t-test; T=-1.88, p=0.06). Depression scores, as measured by GDS, were significantly different between the groups (t-test; T=-2.71, p=7.95X10^-3^). Here, MCI participants reported higher depression severity compared to HC. It is worth noting that despite the statistical trend and significant differences in GAD-7 and GDS respectively, neither of the cohorts would pass the thresholds for anxiety or depression on average. We acknowledge, however, that depression and anxiety are common comorbidities of MCI, which has also been demonstrated in the prior literature [47,48].

### Task-related neural and behavioral features differentiating between MCI and HC

The cognitive tasks in this study were selected for their ability to probe cognitive dysfunction. Specifically, the tasks targeted working memory load (N-Back) and language (Verbal Fluency). There has been evidence of performance deficits in the MCI population in both of the selected cognitive domains [49,50]. Therefore, we extracted behavioral measures of task performance and explored whether they too were able to differentiate between the MCI and HC cohorts.

For N-Back we evaluated both the accuracy (percent correct) and average reaction time (mean time from trial onset to response) for each of the three task conditions, yielding 6 total behavioral measures. Of these, we found that 1-back accuracy (independent t-test: p=1.70×10^-4^) and 2-back accuracy (independent t-test: p=6.71×10^-3^) significantly differentiated between the MCI and HC population (Figure 4a).

**Figure 4.**
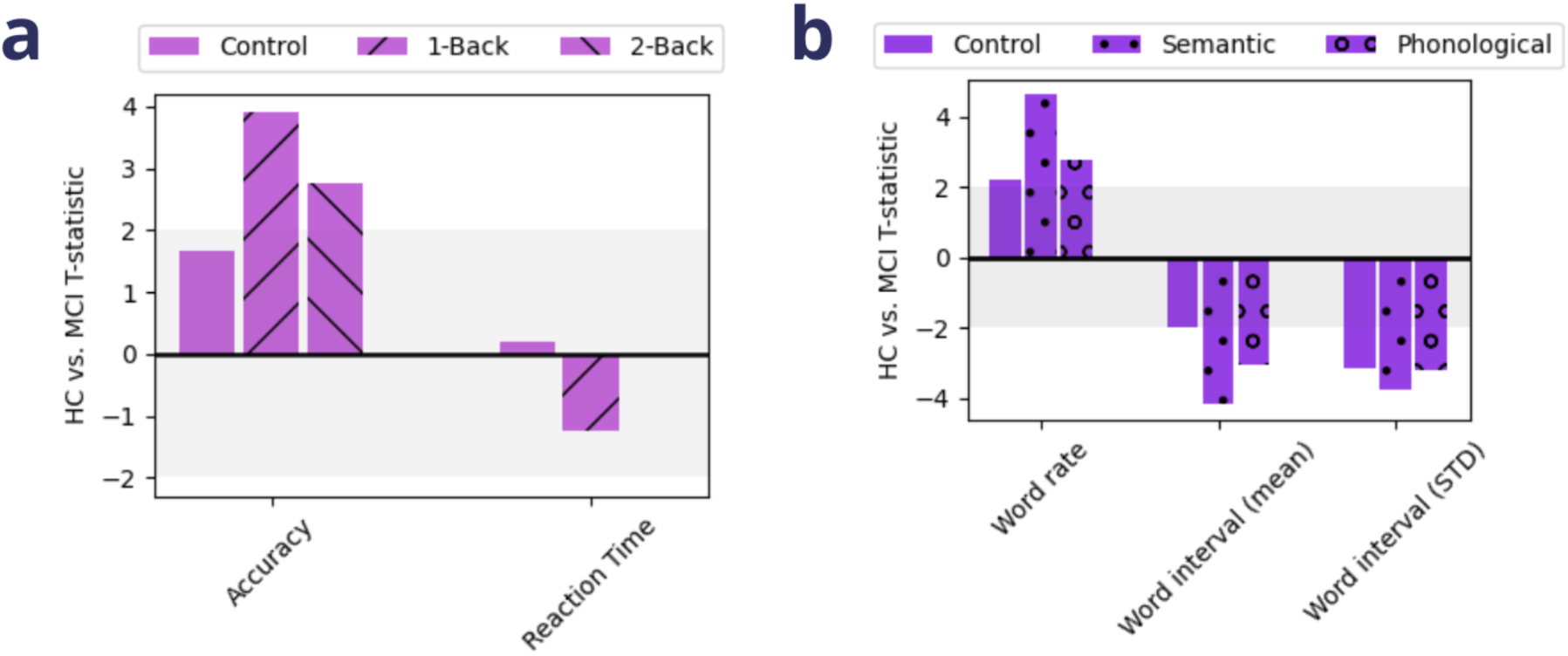
Behavioral performance during cognitive tasks differ between MCI and HC. **a, b.** T-statistics comparing MCI and HC across behavioral measures of task performance within each condition of N-Back **(a)** and Verbal Fluency **(b).** Shaded gray area corresponds to |t-statistics| < 2.

The performance metrics considered for Verbal Fluency were the word rate (rate of word production over the entire block), the average inter-word interval (mean time between words, from first word to last word), and the inter-word variability (standard deviation of time between words) for each of the three task conditions yielding 9 total behavioral measures. Each of these metrics significantly differentiated between the MCI and HC populations, but to varying degrees (Figure 4b). The semantic condition of the task exhibited the strongest differentiability in word rate (p=1.00×10^-5^), inter-word interval (p=7.00×10^-5^), and inter-word variability (p=3.10×10^-4^), followed by the phonological condition (word rate: p=7.06×10^-3^, inter-word interval: p=2.89×10^-3^, inter-word variability: p=1.91×10^-3^), and finally the control condition (word rate: p=0.03, inter-word interval: p=0.048, inter-word variability: p=2.11×10^-3^). Taken together, these differences in task performance reaffirm our task selection and the role of these cognitive domains, working memory and language, in the manifestation of MCI-related impairments.

In order to capture corresponding differences in brain activity explicitly linked to different cognitive demands (of the tasks), hemodynamic response to task conditions was assessed using General Linear Models (GLMs; Methods). These models were applied separately for the Verbal Fluency and N-Back tasks. The resulting GLM test statistics quantified brain activation and deactivation patterns coupled to task conditions over the head. Group GLMs revealed activation and deactivation patterns for the MCI and HC cohorts (Supplement S3, N-Back; Supplement S4, Verbal Fluency). These activations/deactivations were averaged within 35 local regions (corresponding to module locations; Supplement S1) We then compared these values between the MCI and HC populations at each brain region, for both chromophores (HbO/HbR), and for each task condition using independent t-tests.

N-Back showed heightened differentiation in areas linked in the literature to task conditions [51–53] such as regions within the left fronto-temporal cortex (Figure 5a). Notably, this area exhibited reduced HbO during the 0-back condition (lowest working memory load) in HCs compared to MCI participants, possibly reflecting the lower effort required by HCs to complete the task (Figure 5b, top). However, this pattern reversed in the more difficult task conditions. HCs displayed greater neural activity (HbO) in the left fronto-temporal areas, with the strongest response during the highest working memory load (2-back) (Figure 5b, middle, bottom). This increased activity in task-related brain regions may be linked to the significantly better performance of HCs on the 1-back and 2-back tasks. Importantly, neural activity in other brain areas exhibited differences across the two cohorts, indicating that disease-linked differentiability spans the whole head, in line with prior research [52,54].

**Figure 5.**
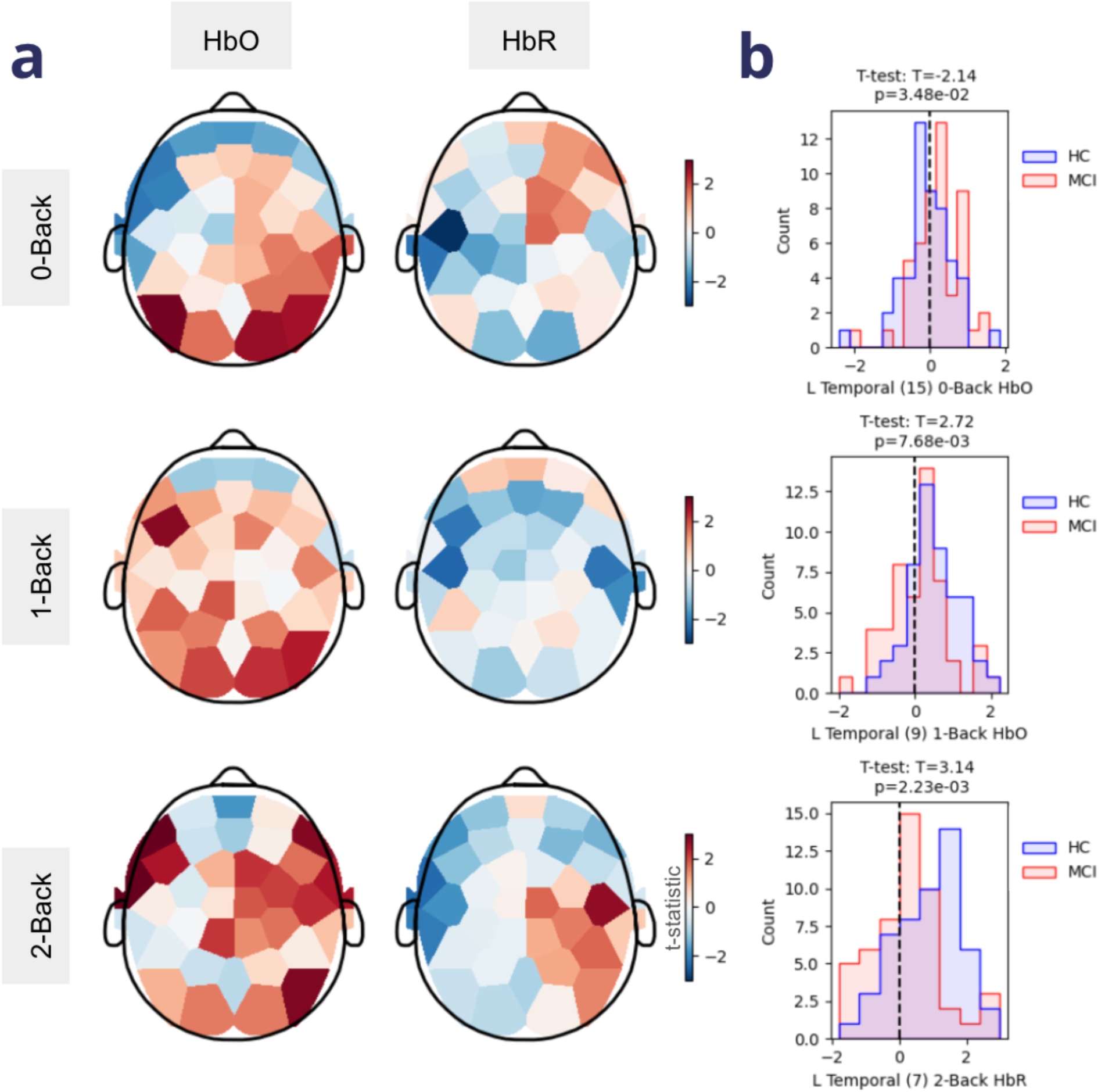
Population differences in hemodynamic responses during the N-Back task. **a.** Shown are t-statistics from independent t-tests between HC with MCI applied to each local area of the GLM-based activation maps. Each row corresponds to a different task condition (top: O-back, middle: 1-back, bottom: 2-back) and the two columns represent HbO/HbR respectively. Warmer colors represent more activation in the HC cohort and cooler colors represent more activation in the MCI cohort, **b.** GLM statistics distributions for representative modules for the HC (blue) and MCI (red) cohorts for each condition (corresponding row). For the location of each module number, see Supplement S1.

Patterns of brain activity linked to conditions of the verbal fluency task not only exhibited group differences in HbO of left fronto-temporal and anterior temporal cortices (Figure 6a, left), areas related to fluency tasks, but also exhibited strong group differences in HbR prominently in occipital regions (Figure 6a, right). While the whole occipital region showed more deactivation in the HC group (compared to the MCI group), the right medial occipital area demonstrated the strongest and most significant difference between the two cohorts during both the control (Figure 6b, top) and semantic conditions of the task. Similarly, the left temporal cortex demonstrated an overall increase in HbO that was strongest and most significant in the inferior medial temporal area (a region subserving categorical word retrieval [55]) during the control and semantic (Figure 6b, middle) conditions of the task. Finally, during the phonological condition the area that most strongly differentiated task-linked brain activity between the HC and MCI cohorts was the left fronto-temporal and prefrontal cortex (Figure 6b, bottom). As with N-back, HCs exhibited stronger activation in task-linked brain areas, while MCI participants showed increased activation (in the case of N-Back) or decreased deactivation (in the case of Verbal Fluency) during the easy conditions of the task. These findings of differential hemodynamic activity to cognitive tasks for MCI vs. HC agree with robust literature findings of similar differences [52,56,57].

**Figure 6.**
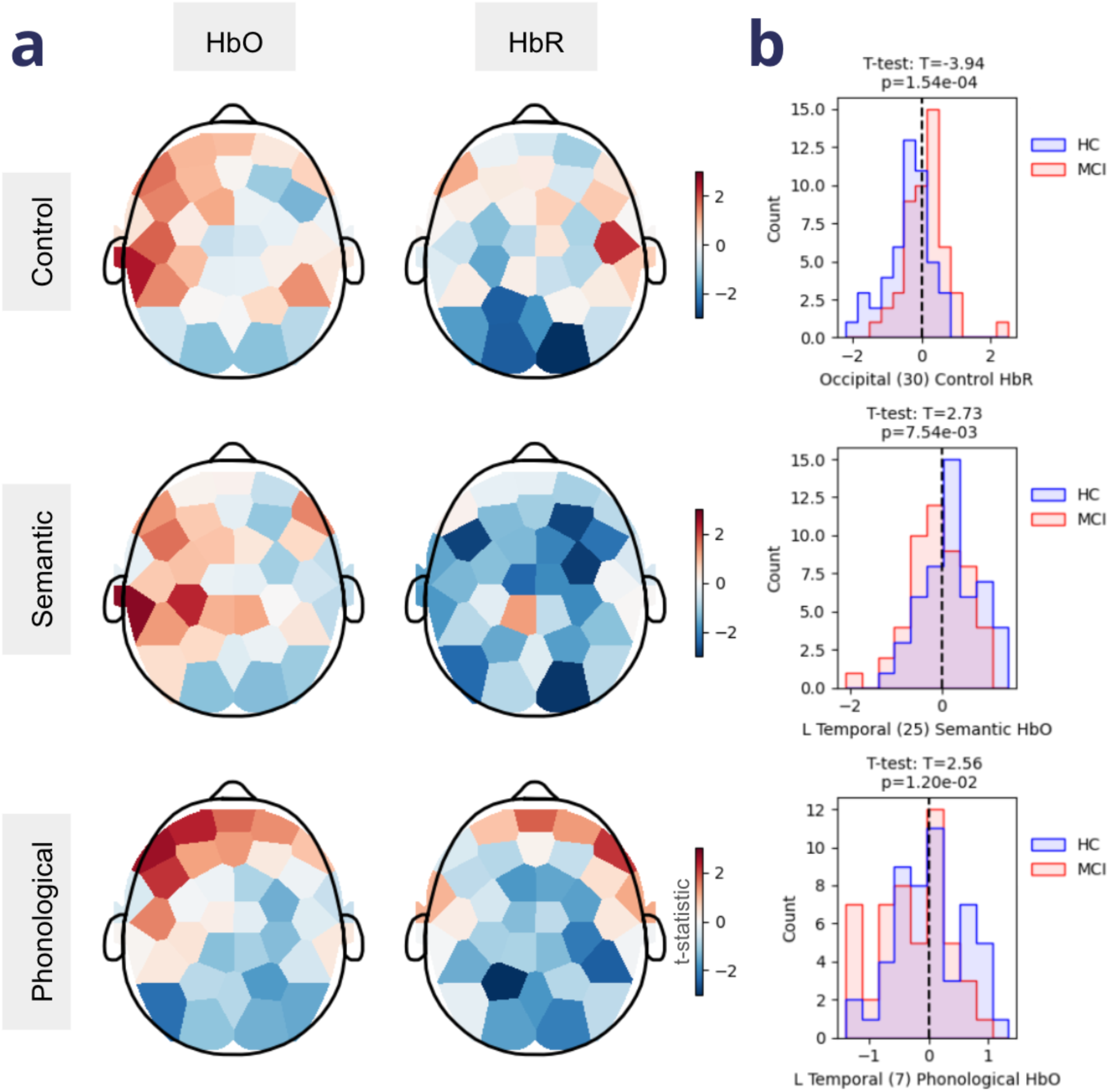
Population differences in hemodynamic responses during the Verbal Fluency task. **a.** Shown are t-statistics from independent t-tests between HC with MCI applied to each region of the GLM-based activation maps. Each row corresponds to a different task condition (top: control, middle: semantic, bottom: phonological) and the two columns represent HbO/HbR respectively. Warmer colors are indicative of more activation in the HC cohort while cooler colors are indicative of more activation in the MCI cohort, b. GLM test statistic distributions for representative modules for the HC (blue) and MCI (red) cohorts for each condition (corresponding row). For the location of each module number, see Supplement S1.

In summary we have shown that not only does task performance discriminate between MCI and HC during specific conditions of the administered cognitive tasks, but so too does the underlying neural activity.

### Machine learning models to detect MCI

While the group-level differences demonstrate potential of the metrics presented here to capture changes in MCI, clinical utility necessitates individual-level diagnostic accuracy. Currently, clinical assessments of MCI consist of subjective questionnaires and cognitive tests. For example, MMSE is often used by clinicians as a screening tool despite its poor sensitivity [7,46]. Indeed, in our MCI population, using a standard cutoff score of <=25 only captures 34% of the MCI population (Figure 3b). In light of this, we asked if using the rich data set at hand in conjunction with ML (Figure 2, Supplement S2), we would be able to detect MCI, and determine which combination of features would afford the highest classification performance. To address this in a systematic way, we built successive machine learning models with different input data and evaluated their performance using standard metrics (Methods).

In the survey-only model, scores from ADCS-ADL-MCI (collected from HC and MCI cohorts) were considered as the sole input, representative of the type of self-report available to clinicians for diagnostic purposes. Similar to previous studies [9], we observed that it is possible to detect MCI using this survey, although the performance of the model is not sufficient to satisfy clinical criteria (Figure 7a, Table 2) [58]. It is worth pointing out that the performance of this model is driven by a high specificity (0.84) or proper labeling of HC individuals, while sensitivity or proper labeling of MCI individuals was rather low (0.62).

**Figure 7.**
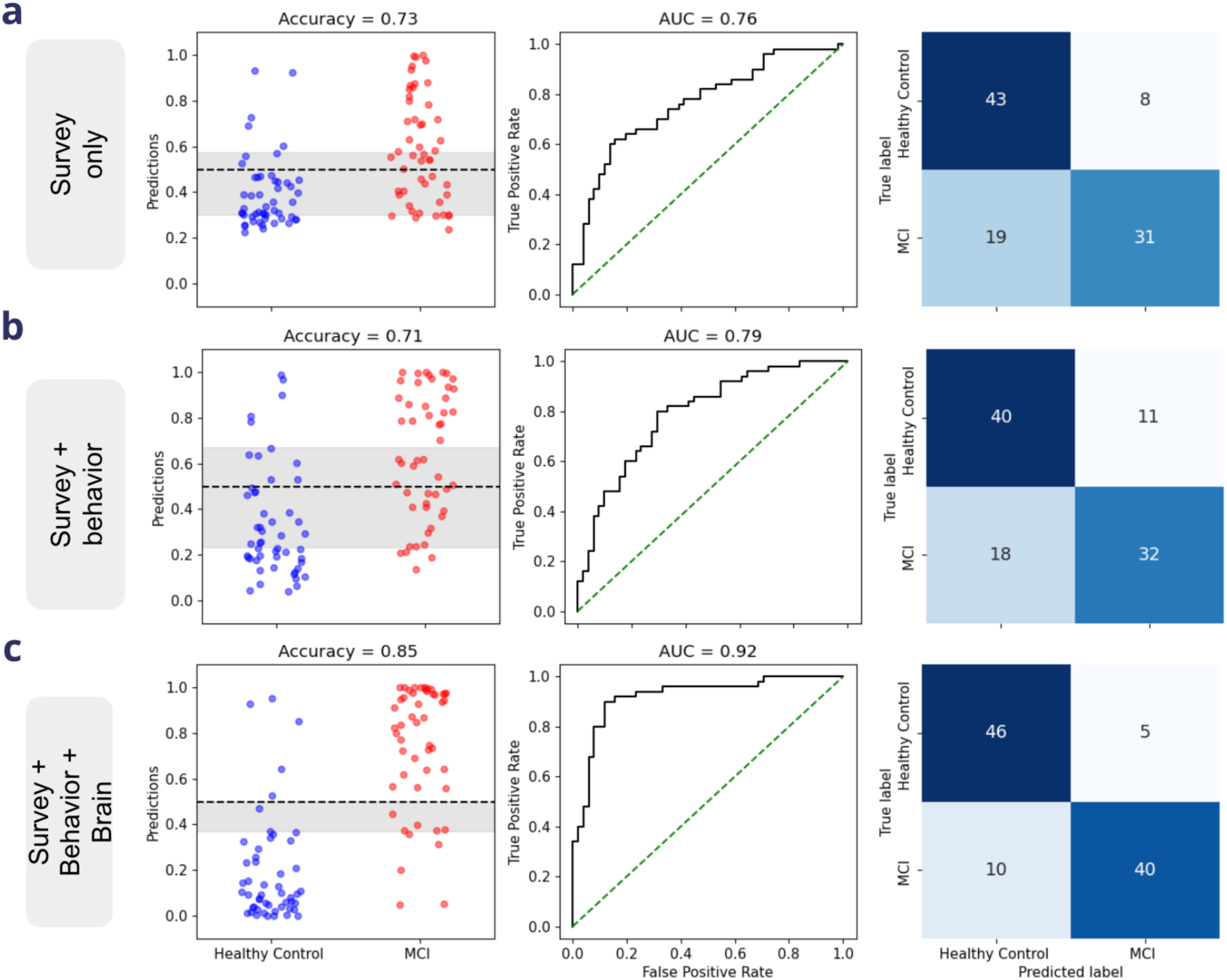
MCI Classification using machine learning underscoring the role of neural features in performance. **a.** Model performance when using only the data from ADCS-ADL-MCI survey score. Note that the model, while good at detecting healthy controls, does not have good sensitivity to MCI. **b.** Combining behavioral metrics from tasks with survey responses did not improve the model performance, **c.** When using survey data as well as both neural and behavioral features from tasks, the model performance was starkly improved. In panels **a-c:** Left) Model prediction raw scores for each group (×-axis). Shaded gray area demonstrate the utility of adapting thresholds to capture an “inconclusive” population. Middle) ROC curve with AUG shown. The diagonal indicates chance level. Right) The confusion matrix when model outputs are binarized.

**Table 2.**
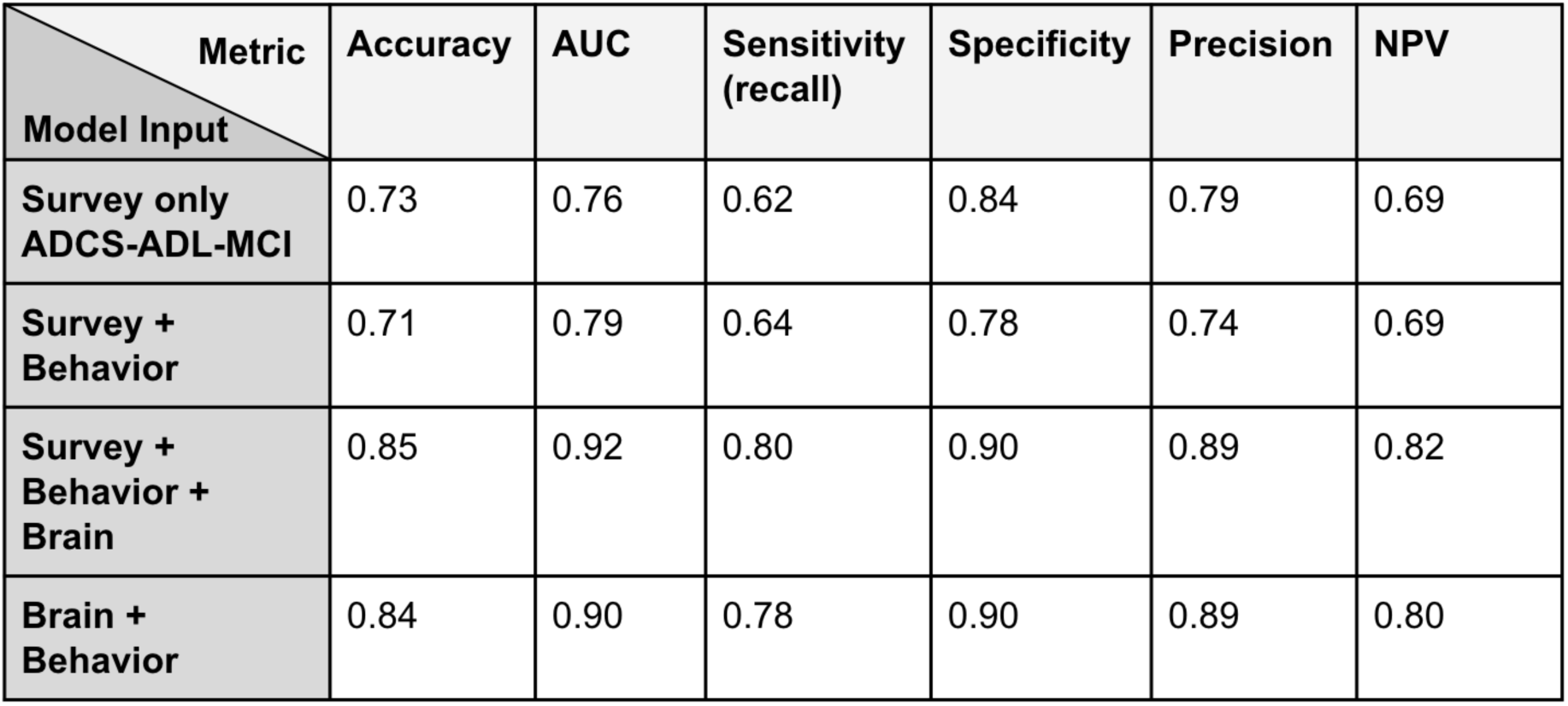
Summary of common performance metrics for the different classification models. Each row indicates a model with the described input data. Behavior data consists of the performance metrics on from both the N-Back and VFT tasks. The brain data consists of GLM features from the two cognitive tasks.

We also performed an additional analysis, beyond the traditional ML metrics, to further quantify the model performance in terms of its predictions (Methods), and highlight ways in which clinicians could use adaptive thresholds to meet screening needs. This method introduced a post-hoc relabeling of participants with inconclusive predictions. Notice the large percentage of participants that would be deemed “Inconclusive” when using only the ADCS-ADL-MCI score (50.50 %).

In addition to survey-based self-reported deficits, we included cognitive performance as input to the model by adding the behavioral metrics from both the verbal fluency and N-back tasks (10 total input features: 9 behavior; 1 survey). Similar task-based measures of cognition may also be available to clinicians. Afterall, behavior on the verbal fluency task has been shown to be useful in detecting MCI [59–62] and, more importantly, some version of this task is administered in diagnostic and screening cognitive tests utilized in clinics (e.g., for both MMSE and MoCA [7,8,30]). This model, with combined behavioral and survey data, performed very similarly to the model with only survey data as inputs (Figure 7b). Here, too, the performance within the HC cohort was much better than the performance of the model within the MCI cohort (i.e., high specificity and low sensitivity respectively). This further suggested that these features by themselves were not sufficient to provide a comprehensive diagnostic power.

Beyond measures currently available to clinicians, we added brain metrics as input to the model. These include GLM-derived measures of task-coupled activity for both chromophores, all brain areas, and all conditions from the Verbal Fluency and N-Back tasks (430 total input features: 420 GLM features; 9 behavior; 1 survey). The performance of this model not only surpassed those of previous models (Figure 7c, Table 2), but also resulted in a sensitivity and specificity that were more closely matched. Notice that the raw scores (Figure 7c, left) have shifted toward more decisive probabilities (e.g., 0 for HC and 1 for MCI). As a result, there are significantly fewer points that fall into the shaded region, representing ‘Inconclusive’ participants (5.95%) in the adaptive decision-making threshold analysis.

The feature selection procedures performed within the nested CV resulted in approximately 100-200 features being passed to the logistic regression model on each outer fold (Supplement S5a). Visualization of the top features in the model (Supplement S5b) revealed that: 1) indeed regularization forced many of the weights towards zero such that the average weight of the top features falls off quickly (exponential decay).; 2) many of the most important features corresponded to the task-related brain areas and behavioral measures that exhibited significant differences between cohorts in our preliminary analyses (e.g., verbal fluency Control HbR Occipital Module 30; 2-Back HbO Left Temporal Module 7; 1-Back accuracy); And 3) neural measures from both tasks, behavioral measures from both tasks, and the ADCS-ADL-MCI survey score all appear within the top 10 features.

Though ADCS-ADL-MCI ranked high in terms of feature importance, we tested whether a model with only task-based features (i.e., neural and behavioral data) could yield satisfactory results without the need to include survey data (429 total input features: 420 GLM; 9 behavior). This is of particular interest given that administering surveys is a time consuming task and self-report questionnaires are vulnerable to misrepresentation of symptomatology, deceit, and interindividual differences in subjective scales. The performance of this model was on par with the model that included ADCS-ADL-MCI survey (Table 2). This suggests that the addition of survey data may not lead to a significant information gain beyond that already available in the task-based features, or at least that the model is able to compensate by increasing the weights of task-based features that manifest some representation of the ADCS-ADL-MCI score.

Taken together, these results suggest that brain-based measures of cognitive function in domains affected by early cognitive impairment could serve as valuable tools for clinicians and primary care doctors. They may also offer a more accurate assessment of cognitive deficits compared to self-reported measures.

## 4. Discussion

In the current study, we sought to examine the ability of detecting MCI using data recorded with the Kernel Flow2 while participants, MCI patients and age-matched HC, performed short commonly-used cognitive tasks. We found that our ML model not only captured expert clinician-based diagnosis of MCI, but also exhibited much higher performance than screening tools such as both the MMSE and self-report ADCS-ADL-MCI surveys. Moreover, the model also shed light on the features necessary for classifying MCI and underscored the importance of both the brain and behavior data from the cognitive tasks.

To classify MCI we used logistic regression with feature selection and regularization, which not only handled a relatively large number of input features, but also provided a clear view of feature importance by examining the model’s weights. The advantage of this approach was the offered transparency and interpretability that can be lacking in more complex models. Consistent with its role as a screening tool, ADCS-ADL-MCI ranked in the top 5 features, though it was prone to false negatives when used in isolation. Furthermore, reflecting the involvement of memory and language dysfunctions in MCI [59,60,63,64], behavioral measures from both tasks were also top features. Highlighting the advantage of using finer-grained quantifications of task performance, it was not word rate, as typically assessed in clinical cognitive tests, but inter-word interval—an indirect measure of word clustering with demonstrated importance in the literature [61,65,66]—that had a more significant role in the model. Finally, we found that neural metrics across both chromophores, all task conditions (including control conditions), and over many regions of the head, especially temporal regions, appeared among the top features. This highlights the necessity of including neural activity recorded over tasks that cover different domains of cognitive impairment, and provides evidence that a similar protocol may be used in the future in lieu of or in addition to standard cognitive assessments.

Another strength of the current approach is that our model labels are based on clinician diagnoses. Our patient cohort was enrolled across several clinical sites with different doctors and clinical protocols. We view this “real world” labeling of our data as a strength, as it allows us to capture something broad and common across diverse protocols for diagnosing patients with MCI. These cognitive assessments usually involve a battery of tests and can take up to 3 hours, therefore introducing more efficient and streamlined procedures, such as short neural recordings, could be beneficial for diagnostic purposes.

Historically neural recordings were cumbersome and/or restricted to hospital settings. Here, we leverage the ease-of-use and portability of the Kernel Flow2 system, which allowed us to carry out the current study at multiple sites. It is possible, nonetheless, that data quality metrics may vary depending on the clinic in which data was collected. While we did not include “data collection site” as a factor in our model and analyses, we acknowledge that this may require additional controls. To ensure homogenous data quality across different locations, we not only monitored incoming data post-hoc but also conducted post-study interviews with individual clinics to receive feedback for further streamlining our protocols, which we plan to integrate into future data collection.

One limitation of our current data collection is potential differences in headset placement on different participants. Although operators are instructed to follow best practices [67] to place the headset according to spatial landmarks on the head, we anticipate variability from participant to participant as well as between placements on a given individual. We have previously shown that within-individual placement variability is minimal [25], and to further alleviate potential differences among participants, we averaged neural activity over larger areas (module-level analyses). Real-time placement aids, such as automated feedback during setup using computer vision can additionally remove this variability.

In the current manuscript we focused on cognitive tasks with behavioral components that are often incorporated into MCI clinical assessments; however, in future analyses we plan to assess both to what extent brain metrics extracted from the resting state recordings (i.e. measures not linked to cognitive demands) can supplement the diagnostic power of our ML models and their stand-alone classification performance. A vast literature on MCI detection using resting state fMRI [12,68] and fNIRS [69,70] data provides evidence that features such as functional connectivity within and across different networks and fALFF may be clinically useful. Another feature that can be extracted from our TD-fNIRS data, absolute oxygenation during the resting state session [18], may offer additional insights into the gradient of cognitive function in healthy controls, MCI, and AD.

Taken together, we showcase the ability of Kernel Flow2 in detecting MCI based on clinician’s diagnosis with high accuracy and performance. Although time-consuming battery of cognitive testing is the current gold standard of MCI diagnosis, we anticipate that neurobehavioral measurements can offer valuable insights into the trajectory of cognitive decline. With adequate testing and wide-spread adoption, these measurements may provide a shorter, more reliable, and more objective standardized replacement for these cognitive tests in the future.

## Data Availability

The datasets used and/or analyzed during the current study are available from the corresponding author on reasonable request.

## 5. Acknowledgements

We gratefully acknowledge the data collection teams at Syrentis Clinical Research and Profound Research, and Dr. Lorrie Bisesi at Syrentis Clinical Research for the operational support. We also thank the participants for their time and contribution to our study.

