## supplementary material for "A functional neuroimaging biomarker of mild cognitive impairment using TD-fNIRS"

### 8. Supplement

##### S1. Kernel Flow headset module numbers throughout the head

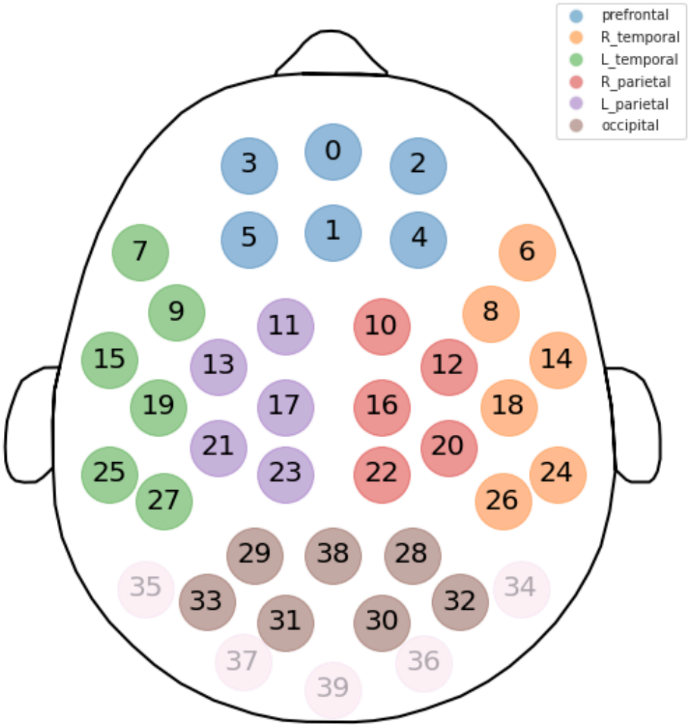


**Figure S1. Topo plot of Kernel Flow2 modules.**
Each circle corresponds to an individual module as indicated by numbers. For this protocol, only 35 of the available 40 modules were used. The used modules are shown here with colored circles and black numbers. Different colors correspond to different regions as described in the manuscript.

#### S2. Machine learning model architecture

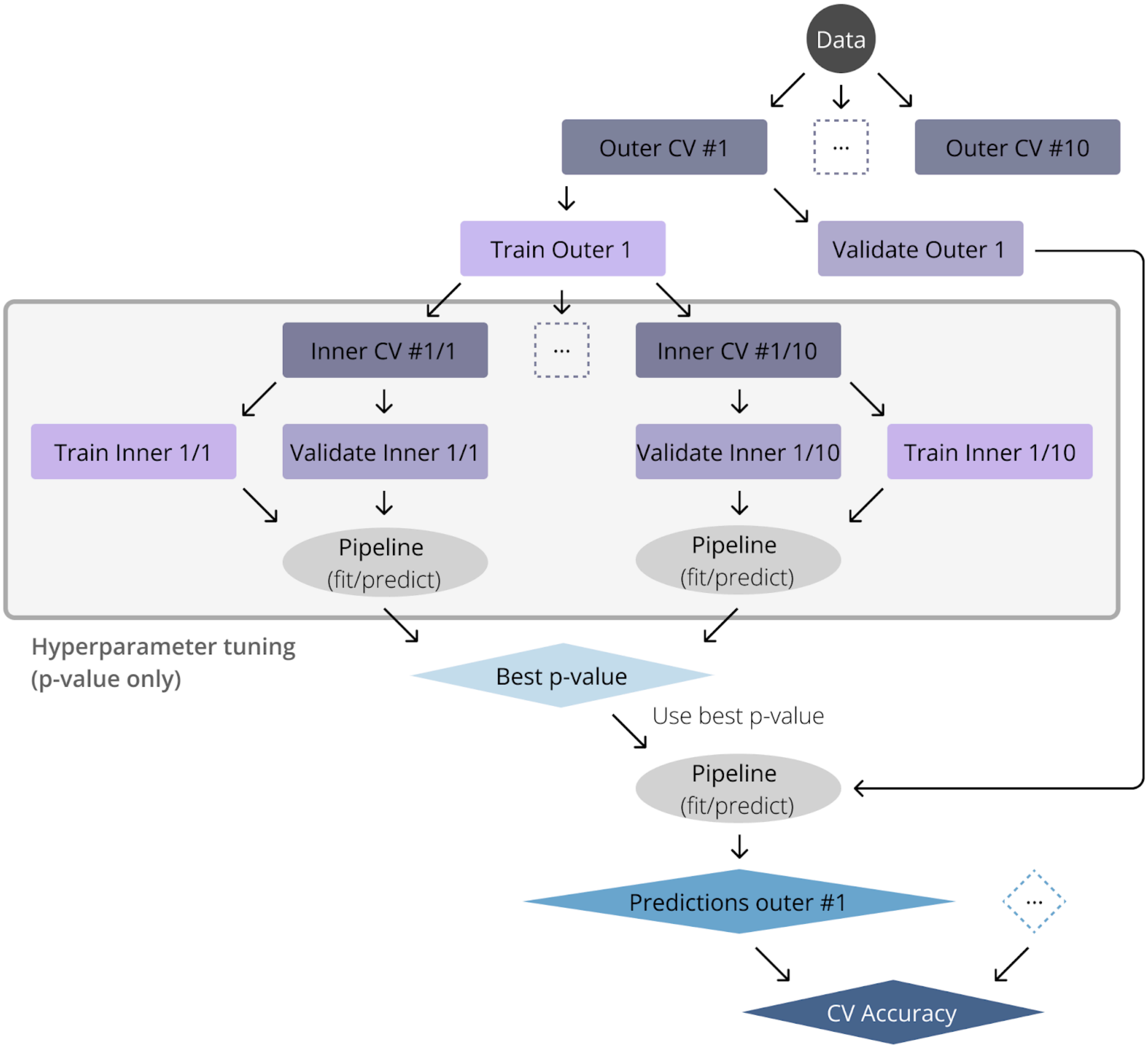


**Figure S2. Additional details about the machine learning model used for MCI classification.** We implemented a nested 10-fold cross validation procedure (N=10 for both inner and outer folds), where the inner fold was used for feature selection and parameter optimization, the outer fold was used to assess model performance on the independent test sets. See Methods for further details.

#### S3. Group Level GLM activations to N-Back task conditions for HC and MCI.

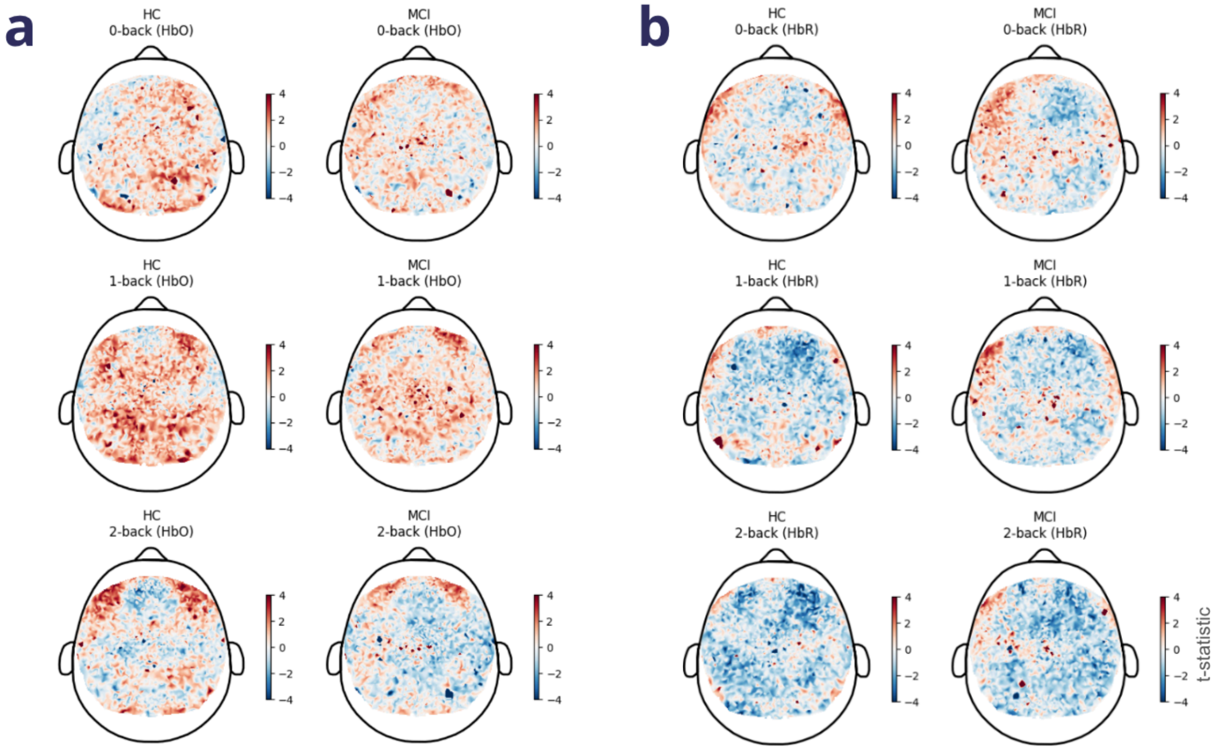


**Figure S3. Brain activation maps during the N-Back task were different between HC and MCI.** Group level GLMs for **a.** HbO (left, HC; right, MCI) and **b.** HbR (left, HC; right, MCI) revealing patterns of activation and deactivation (t-statistics) over the head for each task condition (top, 0-back; middle, 1-back; bottom, 2-back).

**S4. Group Level GLM activations to verbal fluency task conditions for HC and MCI.**

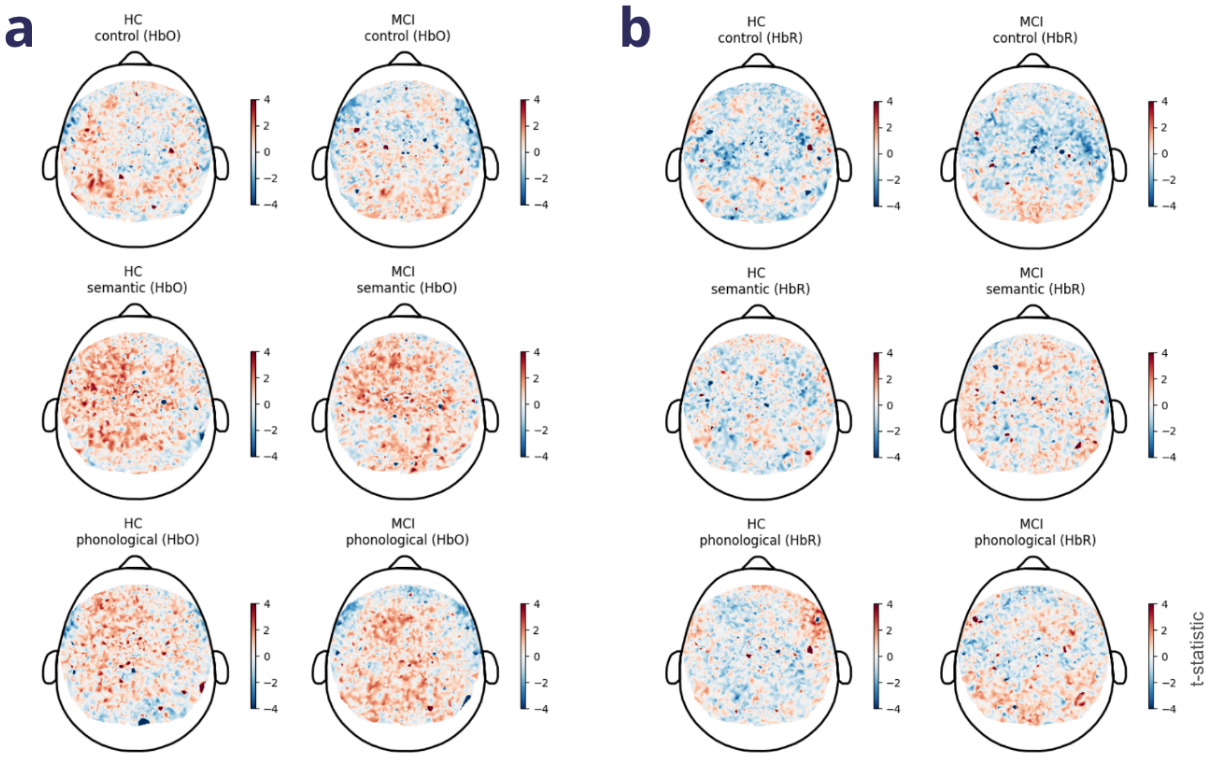


**Figure S4. Brain activation maps during the verbal fluency task were different between HC and MCI.** Group level GLMs for **a.** HbO (left, HC; right, MCI) and **b.** HbR (left, HC; right, MCI) revealing patterns of activation and deactivation (t-statistics) over the head for each task condition (top, control; middle, semantic; bottom, phonological).

#### S5. Important features in MCI classification

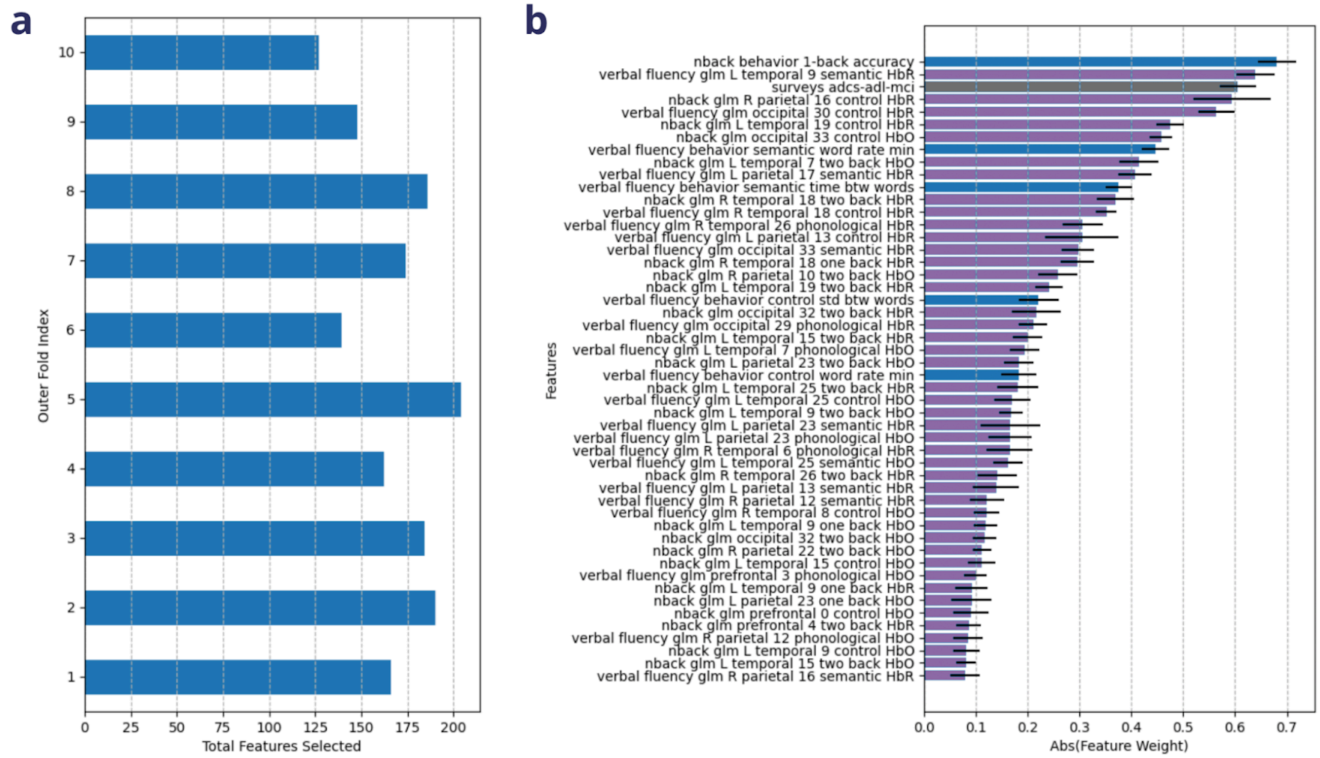


**Figure S5. Further details about input features and their importance in model performance.**
**a.** In the "full" model with input features consisting of GLM features, behavioral and survey data, between 100-200 features were selected in a given outer fold of cross validation (each row represents a different outer fold). **b.** The average weights over the 10 different outer folds (x-axis) for each feature (rows) for the top 50 important features. Features are sorted in descending order by their weights, i.e., how important they are for model performance. Numbers associated with GLM features refer to module number of the headset. Note the quick fall off of weights. Different colors indicate different categories of features: gray represents the ADCS-ADL-MCI survey, blue represents the task behavioral metrics, and purple represents brain features.
